# Effects of Coronavirus Disease 2019 (COVID-19) on Maternal, Perinatal and Neonatal Outcomes: a Systematic Review of 266 Pregnancies

**DOI:** 10.1101/2020.05.02.20088484

**Authors:** Juan Juan, María M. Gil, Zhihui Rong, Yuanzhen Zhang, Huixia Yang, Liona C. Poon

**Affiliations:** Department of Obstetrics and Gynaecology, Peking University First Hospital, Beijing, China; Obstetrics and Gynecology Department, Hospital Universitario de Torrejón, Torrejón de Ardoz, Madrid, Spain; School of Health Sciences, Universidad Francisco de Vitoria (UFV), Pozuelo de Alarcón, Madrid, Spain; Department of Gynaecology and Obstetrics, Tongji Hospital, Tongji Medical College, Huazhong University of Science & Technology; Department of Gynaecology and Obstetrics, Zhongnan Hospital of Wuhan University, Wuhan, China; Clinical Medicine Research Center of Prenatal Diagnosis and Birth Health in Hubei Province, Wuhan, China; Department of Obstetrics and Gynaecology, Prince of Wales Hospital, The Chinese University of Hong Kong, Hong Kong SAR; Harris Birthright Centre, Fetal Medicine Research Institute, King’s College Hospital, King’s College London

**Author notes:** These authors contributed equally to this work (Juan Juan, María M. Gil). Corresponding authors: Prof Huixia Yang, Department of Obstetrics and Gynaecology, Peking University First Hospital, Beijing 100034, China, or, Prof Liona C. Poon, Department of Obstetrics and Gynaecology, Prince of Wales Hospital, The Chinese University of Hong Kong, Hong Kong SAR.

**Keywords:** Coronavirus disease 2019, COVID-19, vertical transmission, spontaneous miscarriage, SARS-CoV-2, spontaneous preterm birth, spontaneous PTB

## Abstract

**Objective:** To perform a systematic review of available published literature on pregnancies affected by COVID-19 to evaluate the effects of COVID-19 on maternal, perinatal and neonatal outcomes.

**Methods:** We performed a systematic review to evaluate the effects of COVID-19 on pregnancy, perinatal and neonatal outcomes. We conducted a comprehensive literature search using PubMed, EMBASE, Cochrane library, China National Knowledge Infrastructure Database and Wan Fang Data until April 20, 2020 (studies were identified through PubMed alert after April 20, 2020). For the research strategy, combinations of the following keywords and MeSH terms were used: SARS-CoV-2, COVID-19, coronavirus disease 2019, pregnancy, gestation, maternal, mothers, vertical transmission, maternal-fetal transmission, intrauterine transmission, neonates, infant, delivery. Eligibility criteria included laboratory-confirmed and/or clinically diagnosed COVID-19, patient was pregnant on admission, availability of clinical characteristics, including maternal, perinatal or neonatal outcomes. Exclusion criteria were unpublished reports, unspecified date and location of the study or suspicion of duplicate reporting, and unreported maternal or perinatal outcomes. No language restrictions were applied.

**Results:** We identified several case-reports and case-series but only 19 studies, including a total of 266 pregnant women with COVID-19, met eligibility criteria and were finally included in the review. In the combined data from seven case-series, the maternal age ranged from 20 to 41 years and the gestational age on admission ranged from 5 to 41 weeks. The most common symptoms at presentation were fever, cough, dyspnea/shortness of breath and fatigue. The rate of severe pneumonia was relatively low, with the majority of the cases requiring intensive care unit admission. Almost all cases from the case-series had positive computer tomography chest findings. There were six and 22 cases that had nucleic-acid testing in vaginal mucus and breast milk samples, respectively, which were negative for SARS-CoV-2. Only a few cases had spontaneous miscarriage or abortion. 177 cases had delivered, of which the majority by Cesarean section. The gestational age at delivery ranged from 28 to 41 weeks. Apgar scores at 1 and 5 minutes ranged from 7 to 10 and 8 to 10, respectively. A few neonates had birthweight less than 2500 grams and over one-third of cases were transferred to neonatal intensive care unit. There was one case each of neonatal asphyxia and neonatal death. There were 113 neonates that had nucleic-acid testing in throat swab, which was negative for SARS-CoV-2. From the case-reports, two maternal deaths among pregnant women with COVID-19 were reported.

**Conclusions:** The clinical characteristics of pregnant women with COVID-19 are similar to those of nonpregnant adults with COVID-19. Currently, there is no evidence that pregnant women with COVID-19 are more prone to develop severe pneumonia, in comparison to nonpregnant patients. The subject of vertical transmission of SARS-CoV-2 remains controversial and more data is needed to investigate this possibility. Most importantly, in order to collect meaningful pregnancy and perinatal outcome data, we urge researchers and investigators to reference previously published cases in their publications and to record such reporting when the data of a case is being entered into a registry or several registries.

**Contribution:** *What are the novel findings of this work?:* Amongst a few cases from the case-series that had qRT-PCR testing in vaginal mucus and breast milks, the results were negative for SARS-CoV-2. Similarly, some of the cases from the case-series had qRT-PCR testing in amniotic fluid, cord blood, neonatal throat swab and neonatal feces, and the results were negative for SARS-CoV-2. Amongst the case-reports, there were two maternal deaths and two neonates tested positive for SARS-CoV-2 at 16 / 24 hours of life.

*What are the clinical implications of this work?:* The subject of vertical transmission of SARS-CoV-2 remains controversial and more data is needed to investigate this possibility. In order to collect meaningful pregnancy and perinatal outcome data, we urge researchers and investigators to reference previously published cases in their publications and to record such reporting when the data of a case is being entered into a registry or several registries.

## INTRODUCTION

With over three million individuals infected,^1^ the global pandemic caused by the severe acute respiratory syndrome coronavirus 2 (SARS-CoV-2), the coronavirus disease 2019 (COVID-19) is a global public health crisis.^2,3^ Most cohort studies have focused on evaluating the effects of COVID-19 on the general population,^2–4^ and there is insufficient data on vulnerable populations, such as pregnant women.

It is recognized that pregnant women are at an increased risk of acquiring viral respiratory infection and developing severe pneumonia due to the physiologic changes in the immune and cardiopulmonary systems.^5,6^ Lessons learned from the two notable coronavirus outbreaks, the severe acute respiratory syndrome coronavirus (SARS-CoV) and the Middle East respiratory syndrome coronavirus (MERS-CoV), suggest that pregnant women are particularly susceptible to adverse outcomes, including the need for endotracheal intubation, admission to an intensive care unit (ICU), renal failure and death.^7–9^ The first study describing the clinical characteristics and investigating the possibility of vertical transmission of SARS-CoV-2 in nine pregnant women with laboratory-confirmed COVID-19, demonstrated that the impact of COVID-19 on pregnant women was similar to that of non-pregnant adults and that there was no evidence of vertical transmission as SARS-CoV-2 was not detected in amniotic fluid, cord blood and neonatal throat swab of six cases.^10^ To date, the largest series reporting on both pregnancy and neonatal outcomes, with a total of 99 COVID-19-infected pregnant women, demonstrated that COVID-19 during pregnancy was not associated with an increased risk of adverse outcomes such as spontaneous preterm birth. Amongst the 100 neonates born to these women, none was infected with SARS-CoV-2.^11^ Based on very scarce data to-date, conflicting evidence from nuclei acid-based testing and antibodies testing in neonates born to mothers with COVID-19 has raised further controversy in relation to the risk of vertical transmission during pregnancy.^12,13^

The objective of this study was to perform a systematic review of available published literature on pregnancies affected by COVID-19 to evaluate the effects of COVID-19 on maternal, perinatal and neonatal outcomes.

## METHODS

### Search strategy

We conducted a comprehensive literature search using PubMed, EMBASE, Cochrane library, China National Knowledge Infrastructure Database and Wan Fang Data until April 20, 2020 (Studies were identified through Pubmed alert after April 20, 2020). For the search strategy, combinations of the following keywords and MeSH terms were used: SARS-CoV-2, COVID-19, coronavirus disease 2019, pregnancy, gestation, maternal, mothers, vertical transmission, maternal-fetal transmission, intrauterine transmission, neonates, infant. The search strategy is provided in Appendix 1. Eligibility criteria included laboratory-confirmed and/or clinically diagnosed COVID-19, patient pregnant on admission, availability of clinical characteristics, including at least one of maternal, perinatal or neonatal outcomes. Exclusion criteria were unpublished reports, unspecified date and location of the study or suspicion of duplicate reporting, and unreported maternal or perinatal outcomes. No language restrictions were applied.

### Selection of the articles

Titles were selected from first screening and abstracts of citations were reviewed by two reviewers independently (J.J. and M.M.G.) to identify all potentially relevant articles. We identified several case-reports and case-series. After the first screening of titles and abstracts, we decided to repeat the screening procedure and exclude case-reports or case-series that include less than 10 cases from China in order to avoid duplication of cases as there have since been several cohort series published. The potentially relevant articles were fully evaluated by the same reviewers. Reference lists of relevant original and review articles were searched for additional reports. Full-text articles were retrieved for further consideration for inclusion. Disagreements were resolved by the opinion of a third party (L.C.P.), who also independently reviewed the final included articles to confirm they met the inclusion criteria. The review was registered in PROSPERO on 23 April 2020,^14^ prior to data extraction (registration number CRD42016032409).

### Data extraction, quality assessment and outcome measures

Two authors (J.J. and M.M.G.) extracted the information (population, outcome, study design and results) from the selected studies. A modification of the Cochrane Public Health Group Data Extraction and Assessment Template,^15^ which was previously piloted by the researchers, was used to tabulate findings of the included articles. Finally, they independently assessed methodological quality of the studies using the Joanna Briggs Institute (JBI) tool for case-series and case-reports.^16^ Publication bias was considered high since all the included studies were either case-series or case-control in design. The quality of this review was validated with the Preferred Reporting Items for Systematic Reviews and Meta-Analyses (PRISMA) tool.^17^

The following information was extracted from the articles: author names, institution and country, study design, sample size, maternal age, gestational age on admission, symptoms on admission, pregnancy complications, gestational age at delivery, mode of delivery, disease severity, laboratory and radiological findings, maternal and neonatal outcomes, samples collection (amniotic fluid, cord blood, placenta, vaginal secretion, urine, feces, breast milk, neonatal pharyngeal swab, neonatal blood, neonatal urine, neonatal feces, neonatal gastric juice). Any evidence of maternal to fetal transmission of SARS-CoV-2 was also recorded. We contacted the authors directly when further clarification on their data was needed, such as overlapping cases in different studies published by the same group or to complete outcome data. Details on correspondence are provided in Supplementary Table 1.

### Statistical analyses

Due to the lack of studies with designs that allow for meta-analysis, we opted to perform a narrative synthesis using the Synthesis Without Meta-analysis (SWiM) reporting guideline (intended to complement the PRISMA guidelines in these cases).^18^ Summary statistics (numbers and ranges) were calculated to show sample distribution, where appropriate.

## RESULTS

Figure 1 summarizes the selection of articles for review. Initial database searches identified 536 records which, after exclusion of duplicates, were screened for the eligibility criteria and included in the current systematic review. Finally, 19 studies, including a total of 266 pregnant women with COVID-19, met the eligibility criteria and were included in the review. There were seven case-series, including 253 pregnant patients, published from March 4, 2020 to April 24, 2020 (Tables 1–6). There were 12 case-reports, including 13 pregnant patients, published from March 25, 2020 to April 18, 2020 (Tables 1–6). From the case-series, all seven studies reported all cases diagnosed during the study period. We recorded data related to maternal and perinatal characteristics, the clinical manifestations of COVID-19 at admission including laboratory testing, treatment received and maternal, perinatal and neonatal outcomes. The majority of the studies originated from China, but cases from Australia, Canada, France, Korea, Iran, Peru, Spain, Sweden, Turkey and United States were also included. We differentiated cases that were laboratory-confirmed from those that were clinically diagnosed but data from the two types of diagnosis were combined for presentation. A laboratory-confirmed case of COVID-19 was defined as a positive result on quantitative reverse-transcriptase-polymerase-chain-reaction (qRT-PCR) assay of maternal pharyngeal swab specimens. It is important to note that at the peak of the COVID-19 outbreak within Hubei province, China, cases with relevant symptoms, significant epidemiological history and typical chest computer tomography (CT) findings were clinically diagnosed as COVID-19, as the viral nucleic acid test was reported to have a false-negative rate of 30%.^19^

**Table 1a:** Characteristics and symptoms of COVID-19 in pregnant women in case-series

| Study | N | Country | Age<br>(Range) | Diagnosis |  | GA on<br>admission<br>(week) | Symptoms |  |  |  |  |  |  |  |
| --- | --- | --- | --- | --- | --- | --- | --- | --- | --- | --- | --- | --- | --- | --- |
|  |  |  |  | Laboratory | Clinical |  | Fever | Cough | Fatigue | Dyspnea/Short<br>ness of breath | Sore<br>throat | Myalgia | Malaise | Diarrhea/GI<br>symptoms |
| Breslin et al<sup>20</sup>, 2020 | 43 | US | 20-39 | 43 | 0 | NR | 14 | 19 | 0 | 7 | 0 | 11 | 0 | 5 |
| Liu et al<sup>36</sup>, 2020 | 41 | China | 22-42 | 16 | 25 | 22-40 | 16 | 15 | 5 | 5 | 0 | 0 | 0 | 0 |
| Liu et al<sup>37</sup>, 2020 | 13 | China | 22-36 | 13 | 0 | NR | 10 | 2 | 4 | 3 | 1 | 0 | 0 | 0 |
| Liu et al<sup>38</sup>, 2020 | 15 | China | 23-40 | 15 | 0 | 12-38 | 13 | 9 | 4 | 1 | 1 | 3 | 0 | 1 |
| Liu et al<sup>39</sup>, 2020 | 19 | China | 26-38 | 10 | 9 | NR | 11 | 5 | 0 | 5 | 0 | 0 | 0 | 2 |
| Wu et al<sup>40</sup>, 2020 | 23 | China | 21-37 | 19 | 4 | 6-40 | 4 | 6 | 0 | 0 | 0 | 0 | 0 | 0 |
| Yan et al<sup>11</sup>, 2020 | 99 | China | 24-41 | 53 | 46 | 5-41<sup>+2</sup> | 50 | 27 | 15 | 10 | 8 | 6 | 0 | 1 |
| Total | 253 |  | 20-41 | 169<br>(66.8%) | 84<br>(33.2%) | 5-41 | 118<br>(46.6%) | 83<br>(32.8%) | 28<br>(11.1%) | 31<br>(12.3%) | 10<br>(3.9%) | 20<br>(7.9%) | 0 | 9<br>(3.6%) |

GA: gestational age; GI: gastrointestinal; US: United States; NR: not reported

**Table 1b:** Characteristics and symptoms of COVID-19 in pregnant women in case-reports

| Study | N | Country | Age<br>(Range) | Diagnosis |  | GA on<br>admission<br>(week) | Symptoms |  |  |  |  |  |  |  |
| --- | --- | --- | --- | --- | --- | --- | --- | --- | --- | --- | --- | --- | --- | --- |
|  |  |  |  | Laboratory | Clinical |  | Fever | Cough | Fatigue | Dyspnea/Short<br>ness of breath | Sore<br>throat | Myalgia | Malaise | Diarrhea/GI<br>symptoms |
| Alonso Diaz et al<sup>41</sup>,<br>2020 | 1 | Spain | 41 | 1 | 0 | 38<sup>+4</sup> | 0 | 1 | 0 | 1 | 0 | 0 | 0 | 0 |
| Alzamora et al<sup>23</sup>, 2020 | 1 | Peru | 41 | 1 | 0 | 33 | 1 | 0 | 1 | 1 | 0 | 0 | 1 | 0 |
| Gidlöf et al<sup>42</sup>, 2020 | 1 | Sweden | 34 | 1 | 0 | 36<sup>+2</sup> | 1 | 0 | 0 | 0 | 0 | 0 | 1 | 0 |
| Gonzalez Romero et<br>al<sup>43</sup>, 2020 | 1 | Spain | 44 | 1 | 0 | 29<sup>+2</sup> | 1 | 1 | 0 | 0 | 1 | 0 | 0 | 0 |
| Iqbal et al<sup>44</sup>, 2020 | 1 | US | 34 | 1 | 0 | 39 | 1 | 1 | 0 | 0 | 0 | 1 | 0 | 0 |
| Kalafat et al<sup>45</sup>, 2020 | 1 | Turkey | 32 | 1 | 0 | 35<sup>+3</sup> | 1 | 1 | 0 | 1 | 0 | 0 | 0 | 0 |
| Karami et al<sup>21</sup>, 2020 | 1 | Iran | 27 | 1 | 0 | 30<sup>+3</sup> | 1 | 1 | 0 | 0 | 0 | 1 | 0 | 0 |
| Lee et al<sup>46</sup>, 2020 | 1 | Korea | 28 | 1 | 0 | 37<sup>+6</sup> | 1 | 1 | 0 | 0 | 1 | 0 | 0 | 0 |
| Lowe et al<sup>47</sup>, 2020 | 1 | Australia | 31 | 1 | 0 | 40<sup>+2</sup> | 1 | 1 | 0 | 0 | 0 | 0 | 0 | 0 |
| Vlachodimitropoulou<br>Koumoutsea et al<sup>48</sup>,<br>2020 | 2 | Canada,<br>France | 23, 40 | 2 | 0 | 35<sup>+5</sup>, 35<sup>+2</sup> | 2 | 2 | 0 | 0 | 0 | 0 | 0 | 0 |
| Zamaniyan et al<sup>22</sup>,<br>2020 | 1 | Iran | 22 | 1 | 0 | 32 | 1 | 1 | 0 | 1 | 0 | 1 | 0 | 1 |
| Zambrano et al<sup>49</sup>, 2020 | 1 | C America | 41 | 1 | 0 | 31 | 1 | 1 | 0 | 0 | 0 | 1 | 0 | 0 |

GA: gestational age; GI: gastrointestinal; US: United States; NR: not reported; C America: Central America

**Table 2a:** Treatment and outcome of COVID-19 in pregnant women in case-series

| Study | N | Treatment |  |  |  |  |  |  | Severe pneumonia | Maternal mortality |
| --- | --- | --- | --- | --- | --- | --- | --- | --- | --- | --- |
|  |  | Antiviral therapy | Antibiotic therapy | Corticosteroid | Hydroxychloroquine | Invasive mechanical ventilation | Noninvasive ventilation | ICU admission |  |  |
| Breslin et al<sup>20</sup>, 2020 | 43 | 0 | 2 | 0 | 2 | 0 | 3 | 2 | 6 | 0 |
| Liu et al<sup>36</sup>, 2020 | 41 | 14 | NR | NR | NR | NR | NR | 0 | 0 | 0 |
| Liu et al<sup>37</sup>, 2020 | 13 | NR | NR | NR | NR | 1 | 0 | 1 | 1 | 0 |
| Liu et al<sup>38</sup>, 2020 | 15 | 11 | 15 | 0 | 0 | 0 | 14 | 0 | 0 | 0 |
| Liu et al<sup>39</sup>, 2020 | 19 | 6 | NR | 0 | NR | NR | NR | NR | NR | 0 |
| Wu et al<sup>40</sup>, 2020 | 23 | NR | NR | NR | NR | NR | NR | NR | 0 | 0 |
| Yan et al<sup>11</sup>, 2020 | 99 | 51 | 94 | 31 | NR | 2 | 4 | 5 | 5 | 0 |
| Total | 253 | 82/217<br>(37.8%) | 111/157<br>(70.7%) | 31/176<br>(17.6%) | 2/58<br>(3.4%) | 3/170<br>(1.8%) | 21/170<br>(12.4%) | 8/211<br>(3.8%) | 12/234<br>(5.1%) | 0 |

ICU: intensive care unit; NR: not reported

**Table 2b:** Treatment and outcome of COVID-19 in pregnant women in case-reports

| Study | N | Treatment |  |  |  |  |  |  | Severe pneumonia | Maternal mortality |
| --- | --- | --- | --- | --- | --- | --- | --- | --- | --- | --- |
|  |  | Antiviral therapy | Antibiotic therapy | Corticosteroid | Hydroxychloroquine | Invasive mechanical ventilation | Noninvasive ventilation | ICU admission |  |  |
| Alonso Diaz et al<sup>41</sup>, 2020 | 1 | NR | NR | NR | NR | 1 | 0 | 1 | 1 | 0 |
| Alzamora et al<sup>23</sup>, 2020 | 1 | 1 | 1 | 1 | 1 | 1 | 0 | 1 | 1 | 0 |
| Gidlof et al<sup>42</sup>, 2020 | 1 | 0 | 1 | 0 | 0 | 0 | 1 | 0 | 0 | 0 |
| Gonzalez Romero et al<sup>43</sup>, 2020 | 1 | 1 | 1 | 1 | 1 | 1 | 0 | 1 | 1 | 0 |
| Iqbal et al<sup>44</sup>, 2020 | 1 | 0 | 0 | 0 | 0 | 0 | 0 | 0 | 0 | 0 |
| Kalafat et al<sup>45</sup>, 2020 | 1 | 1 | 1 | 1 | 1 | 1 | 0 | 1 | 1 | 0 |
| Karami et al<sup>21</sup>, 2020 | 1 | 1 | 1 | 1 | 1 | 1 | 0 | 1 | 1 | 1 |
| Lee et al<sup>46</sup>, 2020 | 1 | 0 | 0 | 0 | 0 | 0 | 0 | 0 | 0 | 0 |
| Lowe et al<sup>47</sup>, 2020 | 1 | 0 | 1 | 0 | 0 | 0 | 0 | 0 | 0 | 0 |
| Vlachodimitropoulou<br>Koumoutsea et al<sup>48</sup>, 2020 | 2 | 0 | 1 | 0 | 0 | NR | NR | 0 | 0 | 0 |
| Zamaniyan et al<sup>22</sup>, 2020 | 1 | 1 | 1 | 1 | 1 | 1 | 0 | 1 | 1 | 1 |
| Zambrano et al<sup>49</sup>, 2020 | 1 | NR | NR | NR | NR | NR | NR | 0 | 0 | 0 |

ICU: intensive care unit; NR: not reported

**Table 3a:** Radiological and laboratory findings of pregnant women with COVID-19 in case-series

| Study | N | CT chest findings |  | Laboratory findings |  |  |  |  |  |  |  |  | Biological samples |  |
| --- | --- | --- | --- | --- | --- | --- | --- | --- | --- | --- | --- | --- | --- | --- |
|  |  | Patchy shadowing<br>or ground-grass<br>opacity | -ve finding | ↓ or ↔<br>leukocyte | ↓ lymphocyte | ↑ CRP | ↑ AST/ALT | ↓ Platelet | ↑ ferritin | ↑ D-dimer | ↑ IgG | ↑ IgM | Vaginal<br>mucus | Breast milk |
| Breslin et al<sup>20</sup>, 2020 | 43 | NR | NR | NR | NR | NR | NR | NR | NR | NR | NR | NR | NR | NR |
| Liu et al<sup>36</sup>, 2020 | 41 | 38 | 3 | 24 | 25 | 27 | NR | NR | NR | NR | NR | NR | NR | NR |
| Liu et al<sup>37</sup>, 2020 | 13 | NR | NR | NR | NR | NR | NR | NR | NR | NR | NR | NR | NR | NR |
| Liu et al<sup>38</sup>, 2020 | 15 | 15 | 0 | NR | 12 | 10 | NR | NR | NR | NR | NR | NR | NR | NR |
| Liu et al<sup>39</sup>, 2020 | 19 | 19 | 0 | NR | NR | NR | NR | NR | NR | NR | NR | NR | NR | 10/10; negative |
| Wu et al<sup>40</sup>, 2020 | 23 | 23 | 0 | NR | NR | NR | NR | NR | NR | NR | NR | NR | NR | NR |
| Yan et al<sup>11</sup>, 2020 | 99 | 88 | 4 | 96 | 42 | 36 | NR | NR | NR | NR | NR | NR | 6/6;<br>negative | 12/12; negative |
| Total | 253 | 183/190<br>(96.3%) | 7/190<br>(3.7%) | 120/140<br>(85.7%) | 79/155<br>(51.0%) | 73/155<br>(47.1%) | NR | NR | NR | NR | NR | NR | - | - |

CT: computer tomography; -ve: negative; CRP: C-reactive protein; AST: Aspartate transaminase; ALT: Alanine transaminase; IgG: immunoglobulin G; IgM: immunoglobulin M; NR: not reported.

**Table 3b:** Radiological and laboratory findings of pregnant women with COVID-19 in case-reports

| Study | N | CT chest findings |  | Laboratory findings |  |  |  |  |  |  |  |  | Biological samples |  |
| --- | --- | --- | --- | --- | --- | --- | --- | --- | --- | --- | --- | --- | --- | --- |
|  |  | Patchy shadowing<br>or ground-grass<br>opacity | -ve finding | ↓ or ↔<br>leukocyte | ↓ lymphocyte | ↑ CRP | ↑ AST/ALT | ↓ Platelet | ↑ ferritin | ↑<br>D-dimer | ↑ IgG | ↑ IgM | Vaginal<br>mucus | Breast milk |
| Alonso Diaz et al<sup>41</sup>,<br>2020 | 1 | NR | NR | NR | NR | NR | NR | NR | NR | NR | NR | NR | NR | NR |
| Alzamora et al<sup>23</sup>, 2020 | 1 | 1 | 0 | 0 | 1 | 1 | 0 | 1 | 1 | 0 | 1 | 1 | NR | NR |
| Gidlof et al<sup>42</sup>, 2020 | 1 | 1 | 0 | 0 | 0 | 1 | 0 | 0 | NR | NR | NR | NR | 1; negative | 1; negative |
| Gonzalez Romero et<br>al<sup>43</sup>, 2020 | 1 | NR | NR | 0 | 1 | 1 | 1 | 0 | NR | 1 | NR | NR | NR | NR |
| Iqbal et al<sup>44</sup>, 2020 | 1 | 1 | 0 | 0 | 1 | 1 | 0 | 0 | NR | 1 | NR | NR | NR | NR |
| Kalafat et al<sup>45</sup>, 2020 | 1 | 1 | 0 | 0 | 1 | NR | NR | 0 | NR | NR | NR | NR | NR | 1; negative |
| Karami et al<sup>21</sup>, 2020 | 1 | 0 | 1 | 0 | 1 | 1 | 1 | 1 | NR | 1 | NR | NR | NR | NR |
| Lee et al<sup>46</sup>, 2020 | 1 | 1 | 0 | 1 | 0 | 1 | 0 | NR | NR | NR | NR | NR | NR | NR |
| Lowe et al<sup>47</sup>, 2020 | 1 | NR | NR | NR | NR | NR | NR | NR | NR | NR | NR | NR | NR | NR |
| Vlachodimitropoulou<br>Koumoutsea et al<sup>48</sup>,<br>2020 | 2 | NR | NR | 2 | 2 | 2 | 2 | 1 | 2 | 2 | NR | NR | NR | NR |
| Zamaniyan et al<sup>22</sup>,<br>2020 | 1 | 1 | 0 | 1 | 1 | 1 | 0 | NR | NR | NR | NR | NR | 1; negative | NR |
| Zambrano et al<sup>49</sup>, 2020 | 1 | NR | NR | NR | NR | NR | NR | NR | NR | NR | NR | NR | NR | NR |

CT: computer tomography; -ve: negative; CRP: C-reactive protein; AST: Aspartate transaminase; ALT: Alanine transaminase; IgG: immunoglobulin G; IgM: immunoglobulin M; NR: not reported.

**Table 4a:**
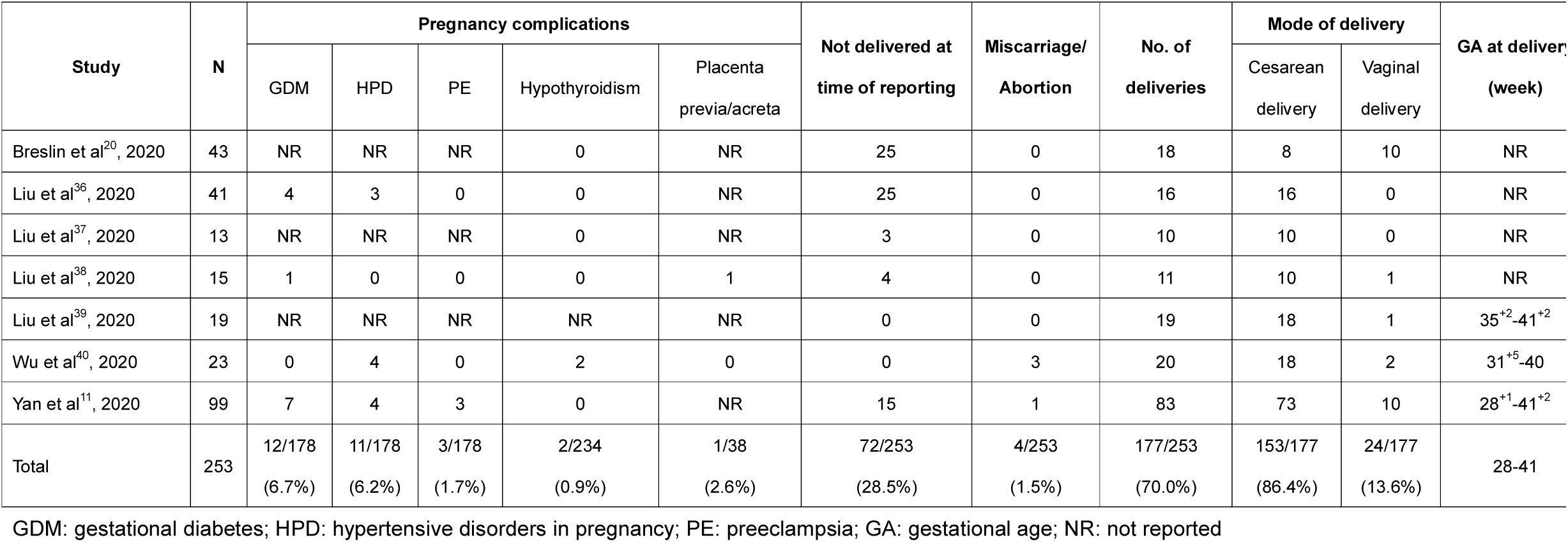
Pregnancy outcomes of COVID-19 in case-series

**Table 4b:** Pregnancy outcomes of COVID-19 in case-reports

| Study | N | Pregnancy complications |  |  |  |  | Not delivered at time of reporting | No. of deliveries | Mode of delivery |  | GA at delivery (week) |
| --- | --- | --- | --- | --- | --- | --- | --- | --- | --- | --- | --- |
|  |  | GDM | HPD | PE | Hypothyroidism | Placenta previa/acreta |  |  | Cesarean delivery | Vaginal delivery |  |
| Alonso Diaz et al<sup>41</sup>, 2020 | 1 | 0 | 1 | 1 | NR | 0 | 0 | 1 | 1 | 0 | 38<sup>+4</sup> |
| Alzamora et al<sup>23</sup>, 2020 | 1 | NR | 0 | 0 | 0 | 0 | 0 | 1 | 1 | 0 | 33<sup>+3</sup> |
| Gidlof et al<sup>42</sup>, 2020 | 1 | 1 | 1 | 1 | 0 | 0 | 0 | 1 | 1 | 0 | NR |
| Gonzalez Romero et al<sup>43</sup>, 2020 | 1 | 0 | 0 | 0 | 0 | 0 | 0 | 1 | 1 | 0 | 29 |
| Iqbal et al<sup>44</sup>, 2020 | 1 | 0 | 0 | 0 | NR | 0 | 0 | 1 | 0 | 1 | 39 |
| Kalafat et al<sup>45</sup>, 2020 | 1 | NR | 0 | 0 | NR | NR | 0 | 1 | 1 | 0 | NR |
| Karami et al<sup>21</sup>, 2020 | 1 | 0 | 0 | 0 | 0 | 0 | 0 | 1 | 0 | 1 | 30<sup>+3</sup> |
| Lee et al<sup>46</sup>, 2020 | 1 | NR | 0 | 0 | 0 | 0 | 0 | 1 | 1 | 0 | 37<sup>+6</sup> |
| Lowe et al<sup>47</sup>, 2020 | 1 | 0 | 0 | 0 | 0 | 0 | 0 | 1 | 0 | 1 | 40<sup>+3</sup> |
| Vlachodimitropoulou Koumoutsea et al<sup>48</sup>, 2020 | 2 | 1 | 0 | 0 | 0 | 0 | 0 | 2 | 2 | 0 | 36, 35<sup>+5</sup> |
| Zamaniyan et al<sup>22</sup>, 2020 | 1 | 0 | 0 | 0 | 0 | 0 | 0 | 1 | 1 | 0 | 32<sup>+4</sup> |
| Zambrano et al<sup>49</sup>, 2020 | 1 | 0 | 1 | 0 | 1 | 0 | 1 | 0 | 0 | 0 | NR |

GDM: gestational diabetes; HPD: hypertensive disorders in pregnancy; PE: preeclampsia; GA: gestational age; NR: not reported

**Table 5a:** Neonatal outcomes of COVID-19 in case-series

| Study | N<br>(neonates) | Apgar score, 1<br>min (Range) | Apgar score,<br>5 min<br>(Range) | Birthweight<br><2500g | Transferred<br>to NICU | Neonatal<br>asphyxia | Neonatal medical complications |  |  |  |  |  |  | Neonatal<br>mortality |
| --- | --- | --- | --- | --- | --- | --- | --- | --- | --- | --- | --- | --- | --- | --- |
|  |  |  |  |  |  |  | Neonatal<br>pneumonia | Shortness<br>of breath | Vomiting | Fever | Cough | Respiratory<br>tract<br>symptoms | Dyspnea |  |
| Breslin et al<sup>20</sup>, 2020 | 18 | 7-10 | 9-10 | NR | 3 | NR | 0 | 0 | 0 | 0 | 0 | 1 | 0 | 0 |
| Liu et al<sup>36</sup>, 2020 | 16 | NR | NR | NR | NR | 0 | NR | NR | NR | NR | NR | NR | NR | 0 |
| Liu et al<sup>37</sup>, 2020 | 10 | 10 | NR | NR | 1 | 1 | NR | NR | NR | NR | NR | NR | NR | 1 |
| Liu et al<sup>38</sup>, 2020 | 11 | 8 | 9 | NR | NR | 0 | NR | NR | NR | NR | NR | NR | NR | 0 |
| Liu et al<sup>39</sup>, 2020 | 19 | 8 | 9 | 0 | 0 | 0 | 0 | 0 | 0 | 0 | 0 | 0 | 0 | 0 |
| Wu et al<sup>40</sup>, 2020 | 21* | NR | 9-10 | NR | NR | 0 | NR | NR | NR | NR | NR | NR | NR | 0 |
| Yan et al<sup>11</sup>, 2020 | 84* | 7-10 | 8-10 | 8 | 42 | 0 | NR | NR | NR | NR | NR | NR | NR | 0 |
| Total | 179 | 7-10 | 8-10 | 8/103<br>(7.8%) | 46/116<br>(39.7%) | 1/161<br>(0.6%) | 0/37 | 0/37 | 0/37 | 0/37 | 0/37 | 1/37<br>(2.7%) | 0/37 | 1/179<br>(0.6%) |

\*Including 1 pregnancy with twin. NR: not reported.

**Table 5b:**
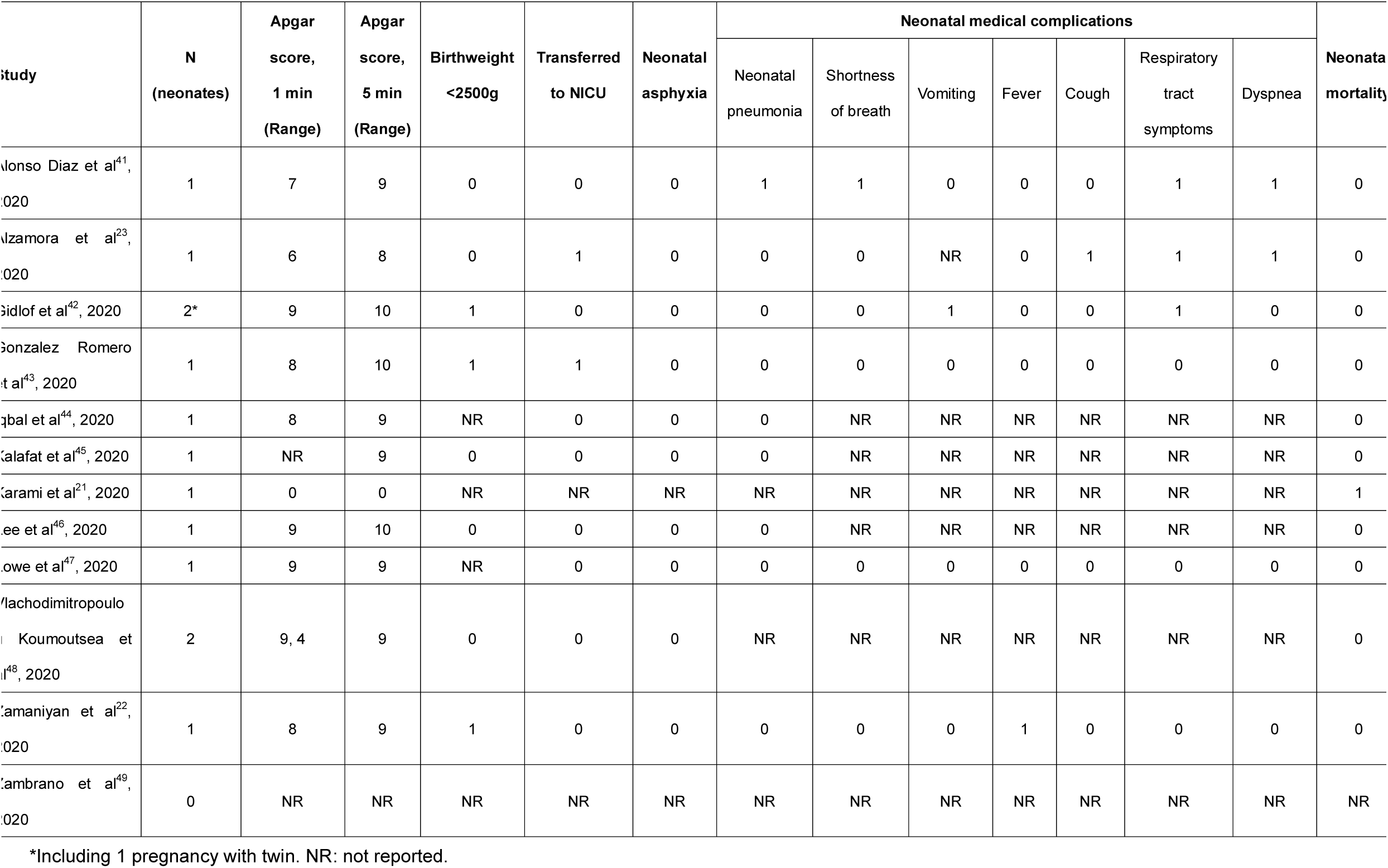
Neonatal outcomes of COVID-19 in case-reports

**Table 6a:** Laboratory and samples of neonates in case-series

| Study | N<br>(neonates) | Laboratory |  |  |  |  |  | Samples |  |  |  |  |  |  |
| --- | --- | --- | --- | --- | --- | --- | --- | --- | --- | --- | --- | --- | --- | --- |
|  |  | ↑ WBC | ↓ lymphocyte | ↓ Platelet | ↑ CRP | ↑ IgG | ↑ IgM | Amniotic fluid | Cord blood | Placenta | Throat swab | Feces | Urine | Gastric juice |
| Breslin et al<sup>20</sup>,<br>2020 | 18 | NR | NR | NR | NR | NR | NR | NR | NR | NR | 18; negative | NR | NR | NR |
| Liu et al<sup>36</sup>,<br>2020 | 16 | NR | NR | NR | NR | NR | NR | NR | NR | NR | NR | NR | NR | NR |
| Liu et al<sup>37</sup>,<br>2020 | 10 | NR | NR | NR | NR | NR | NR | NR | NR | NR | NR | NR | NR | NR |
| Liu et al<sup>38</sup>,<br>2020 | 11 | NR | NR | NR | NR | NR | NR | NR | NR | NR | NR | NR | NR | NR |
| Liu et al<sup>39</sup>,<br>2020 | 19 | 4 | 0 | 0 | 2 | NR | NR | 19; negative | 19; negative | NR | 19; negative | 19; negative | 19; negative | 19; negative |
| Wu et al<sup>40</sup>,<br>2020 | 21* | NR | NR | NR | NR | NR | NR | NR | NR | NR | 4; negative | NR | NR | NR |
| Yan et al<sup>11</sup>,<br>2020 | 84* | NR | NR | NR | NR | NR | NR | 10; negative | 10; negative | NR | 72; negative | NR | NR | NR |
| Total | 179 | 4/19<br>(21.1%) | 0/19 | 0/19 | 2/19<br>(10.5%) | NR | NR | - | - | - | - | - | - | - |

\*Including 1 pregnancy with twin. NR: not reported. CRP: C-reactive protein; IgG: immunoglobulin G; IgM: immunoglobulin M; NR: not reported.

**Table 6b:** Laboratory and samples of neonates in case-reports

| Study | N<br>(neonates) | Laboratory |  |  |  |  |  | Samples |  |  |  |  |  |  |
| --- | --- | --- | --- | --- | --- | --- | --- | --- | --- | --- | --- | --- | --- | --- |
|  |  | ↑ WBC | ↓<br>lymphocyte | ↓<br>Platelet | ↑ CRP | ↑ IgG | ↑ IgM | Amniotic<br>fluid | Cord blood | Placenta | Throat<br>swab | Feces | Urine | Gastric<br>juice |
| Alonso Diaz et al<sup>41</sup>, 2020 | 1 | NR | NR | NR | 0 | NR | NR | NR | NR | NR | 1; negative | NR | NR | NR |
| Alzamora et al<sup>23</sup>, 2020 | 1 | 0 | 0 | 0 | 0 | 0 | 0 | NR | NR | NR | 1; positive | NR | NR | NR |
| Gidlof et al<sup>42</sup>, 2020 | 2* | NR | NR | NR | NR | NR | NR | NR | NR | NR | 2; negative | NR | NR | NR |
| Gonzalez Romero et al<sup>43</sup>,<br>2020 | 1 | NR | NR | NR | NR | NR | NR | NR | NR | NR | 1; negative | NR | NR | NR |
| Iqbal et al<sup>44</sup>, 2020 | 1 | NR | NR | NR | NR | NR | NR | NR | NR | NR | 1; negative | NR | NR | NR |
| Kalafat et al<sup>45</sup>, 2020 | 1 | NR | NR | NR | NR | NR | NR | NR | 1; negative | 1; negative | 1; negative | NR | NR | NR |
| Karami et al<sup>21</sup>, 2020 | 1 | NR | NR | NR | NR | NR | NR | NR | NR | NR | NR | NR | NR | NR |
| Lee et al<sup>46</sup>, 2020 | 1 | NR | NR | NR | NR | NR | NR | 1; negative | 1; negative | 1; negative | 1; negative | NR | NR | NR |
| Lowe et al<sup>47</sup>, 2020 | 1 | NR | NR | NR | NR | NR | NR | NR | NR | NR | 1; negative | NR | NR | NR |
| Vlachodimitropoulou<br>Koumoutsea et al<sup>48</sup>, 2020 | 2 | NR | NR | NR | NR | NR | NR | NR | NR | NR | NR | NR | NR | NR |
| Zamaniyan et al<sup>22</sup>, 2020 | 1 | NR | NR | NR | NR | NR | NR | 1; positive | 1; negative | NR | 1; negative | NR | NR | NR |
| Zambrano et al<sup>49</sup>, 2020 | 0 | NR | NR | NR | NR | NR | NR | NR | NR | NR | NR | NR | NR | NR |

\*Including 1 pregnancy with twin. NR: not reported. CRP: C-reactive protein; IgG: immunoglobulin G; IgM: immunoglobulin M; NR: not reported.

Reasons for exclusion are provided in Supplementary Table 2. Quality assessment of the included studies is reported in Supplementary Tables 3 and 4.

### Clinical characteristics

In the combined data from consecutive case-series, the maternal age ranged from 20 to 41 years and the gestational age on admission ranged from 5 to 41 weeks. Two-thirds of the data derived from laboratory-confirmed COVID-19 cases. The most common symptoms at presentation were fever, cough, dyspnea/shortness of breath and fatigue. Based on the case-series by Yan et al and Breslin et al, up to one-third of COVID-19-infected pregnant patients were asymptomatic on admission.^11,20^ The rate of severe pneumonia was relatively low, with the majority of the cases requiring ICU admission, of which only a few cases received invasive mechanical ventilation. The majority of cases received antibiotic therapy, whilst the minority of cases received antiviral therapy and corticosteroids. Few cases received hydroxychloroquine. There were no cases of maternal death from the case-series.

Almost all cases from the case-series had positive CT chest findings. On admission, the majority of cases had normal or low leucocytes, whilst half had lymphocytopenia and increased C-reactive protein (CRP). Of note, there were six and 22 cases that had nucleic-acid testing in vaginal mucus and breast milk samples, respectively, which were negative for SARS-CoV-2.

From the case-reports, two maternal deaths among pregnant women with COVID-19 were reported by Karami et al. and Zamaniyan et al. from Iran.^21,22^ The first case was a 27-year-old 30 weeks pregnant woman who complained of fever, cough, and myalgia for 3 days. Her admission laboratory tests showed leukopenia and thrombocytopenia, accompanied with elevated CRP and lactate dehydrogenase levels [R]. Soon after admission, her temperature was noted at 40°C and her respiratory rate was 55 per minute, accompanied by suprasternal and intercostal retraction. Immediate blood tests showed metabolic alkalosis while the patient was under non-invasive ventilation. She was eventually intubated for mechanical ventilation due to worsening acute respiratory distress syndrome (ARDS) based on clinical and radiological findings. On day 2 of admission, the patient had spontaneous onset of labor and delivered a cyanotic neonate vaginally with no signs of life that did not respond to neonatal cardiopulmonary resuscitation. One day after birth, the mother developed multi-organ failure (ARDS, acute kidney injury, and septic shock) and died.^21^ The second case was a 22-year-old 32 weeks pregnant woman with dyspnea, myalgia, anorexia, nausea, non-productive cough and fever for 4 days. On admission, she was treated with Azithromycin, Ceftriaxone, Kaletra, Tamiflu and hydroxychloroquine. Based on the CT chest findings, lymphopenia, worsening pneumonia symptoms, and an unfavorable cervix for induction, a Cesarean delivery was indicated to terminate the pregnancy at 33 weeks’ gestation. A preterm female infant, weighing 2350 grams, was delivered uneventfully; with Apgar scores of 8 and 9 at 1 and 5 minutes, respectively. The amniotic fluid tested positive for SARS-CoV-2 by qRT-PCR. The immediate post-delivery nasal and throat swabs of the newborn tested negative for SARS-CoV-2, however, repeat testing 24 hours later became positive. Such results raised the possibility of vertical transmission. The mother underwent peritoneal dialysis due to ARDS on day 4 and 6 postpartum, and required intubation and mechanical ventilation due to sudden oxygen desaturation to 70% on day 10 postpartum. She developed emphysema after intubation that resolved spontaneously on day 12 postpartum, however, her condition deteriorated dramatically and she died on day 15 postpartum.^22^

### Pregnancy and Neonatal Outcomes

From the case-series, the rates of gestational diabetes, hypertensive disorders of pregnancy and preeclampsia did not appear to be higher than in pregnant women without COVID-19. There were only a few cases with hypothyroidism and placenta previa/acreta. Nearly one-third of the cases (72/253) were not delivered at the time of reporting. A few cases had spontaneous miscarriage or abortion. In the remaining 177 cases, including two with twin pregnancy, the majority had a Cesarean delivery. The gestational age at delivery ranged from 28 to 41 weeks. The Apgar scores at 1 and 5 minutes ranged from 7 to 10 and 8 to 10, respectively. A few neonates had birthweight less than 2500 grams and over one-third of cases were transferred to neonatal intensive care unit (NICU). There was one case each of neonatal asphyxia and neonatal death. For the neonates, a few cases had increased white blood cells and CRP. No cases had lymphocytopenia and thrombocytopenia. Of note, there were 29, 29, 113, 19, 19 and 19 cases that had nucleic-acid testing in amniotic fluid, cord blood, neonatal throat swab, neonatal feces, neonatal urine and neonatal gastric juice samples, respectively, which were negative for SARS-CoV-2.

There was a case-report demonstrating that the neonate throat swab tested positive for SARS-CoV-2.^23^ The mother was a 41-year-old woman with pre-existing diabetes mellitus and significant COVID-19 exposure from immediate family members. She presented at 33 weeks’ gestation with a 4-day history of malaise, fatigue, low-grade fever, and progressive shortness of breath. The nasopharyngeal swab of the patient was positive for SARS-CoV-2. The patient developed severe respiratory failure requiring mechanical ventilation on day 5 of disease onset. She was started on azithromycin, hydroxychloroquine, meropenem, vancomycin, and oseltamivir. The patient underwent a preterm Cesarean delivery due to compromised respiratory status. Neonatal isolation was implemented immediately after birth, without delayed cord clamping or skin-to-skin contact. The neonate weighed 2970 grams, with Apgar’s scores of 6 and 8 at 1 and 5 minutes, respectively. The neonate was not exposed to family members. Breastfeeding was not initiated. The neonate was placed in the NICU with no other COVID-19 cases, as this was the first pediatric case at the institution. Chest X-ray showed no abnormalities. At 16 hours after delivery, the neonatal nasopharyngeal swab tested positive for SARS-CoV-2 by RT-PCR, which was repeated 48 hours later that remained positive. However, anti-SARS-CoV-2 IgM and IgG were negative at birth. The possibility of postpartum neonatal infection cannot be completely excluded because of the delay in testing. The newborn required ventilatory support for 12 hours, after which he was extubated and placed on continuous positive airway pressure, with favorable outcome.

## DISCUSSION

This systematic review has demonstrated that, first, the clinical characteristics of these patients with COVID-19 pneumonia during pregnancy are similar to those of nonpregnant adults with COVID-19;^24,25^ second, the most common symptoms reported are fever, cough, dyspnea/shortness of breath and fatigue; third, on admission, most cases have patchy shadowing or ground-glass opacity on CT of the chest, and that normal or low leukocyte, lymphocytopenia and raised CRP are the most common laboratory findings observed in COVID-19-infected pregnant patients; fourth, the rate of severe COVID-19 pneumonia is relatively low but there are two reported maternal deaths, as of April 23, 2020; fifth, COVID-19 does not appear to increase the risk of adverse pregnancy outcomes such as preeclampsia; sixth, only a few pregnancies have resulted in a spontaneous miscarriage or abortion; seventh, of those who have delivered, the gestational age at delivery ranged from 28 to 41 weeks and the majority of cases have had Cesarean delivery; and eighth, in the case-series there have been no reported cases of neonates tested positive for SARS-CoV-2, however, in the case-reports there has been one case each with positive SARS-CoV-2 in amniotic fluid and neonatal throat swab.

At present, there is much controversy relating to the possibility of vertical mother-to-baby transmission of SARS-CoV-2. In the two earlier studies, with a combined total of ten pregnant women with COVID-19 in the third trimester, amniotic fluid, cord blood and neonatal throat-swab samples tested negative for SARS-CoV-2, suggesting there was no evidence of vertical transmission in women who developed COVID-19 pneumonia in late pregnancy.^10,26^ In another series, a neonate born to a pregnant woman with COVID-19 that tested positive for SARS-CoV-2 in the pharyngeal swab sample 36 hours after birth, it was subsequently confirmed that qRT-PCR testing of the placenta and cord blood was negative for SARS-CoV-2, suggesting that intrauterine vertical transmission might not have occurred.^27,28^

Two research letters^12,13^ have suggested the possibility of vertical transmission of SARS-CoV-2 based on the presence of IgM antibodies in blood drawn from three neonates born to COVID-19-infected mothers. However, for all three, the respiratory samples tested negative for SARS-CoV-2. Furthermore, one neonate had repeated testing for SARS-CoV-2 IgG and IgM antibodies, and the observed rapid decline of IgG levels within 14 days, along with a decline in IgM antibodies, strongly suggested that neonatal SARS-CoV-2 IgG antibodies were derived transplacentally from the mother, and not actively induced by the presumed neonatal infection.

The findings of the case of maternal death reported by Zamaniyan et al,^22^ where amniotic fluid and neonatal nasal and throat swabs tested positive for SARS-CoV-2, and the case-report by Alzamora et al,^23^ where neonatal throat swab tested positive for SARS-CoV-2, have again raised the possibility of vertical transmission. The authors of the former case-report could not measure specific antibodies in the neonate, which might have provided supplementary evidence for vertical transmission of SARS-CoV-2. This case is of particular interest because the pregnant patient might have had virulent COVID-19, which was not immediately apparent prior to delivery as her clinical course only dramatically deteriorated after delivery. There appear at least two ways through which SARS-CoV-2 can cause intrauterine infection by vertical transmission. Angiotensin-converting enzyme 2 (ACE2), which has recently been indicated as the putative surface receptor of sensitive cells for SARS-CoV-2,^29^ has been shown to be expressed in human placentas.^30^ This opens up the possibility for SARS-CoV-2 to spread transplacentally through ACE2. Specifically, the viral surface spike glycoprotein (S protein) of SARS-CoV-2 is cleaved by transmembrane protease serine 2 (TMPRSS2) to facilitate efficiency of entry and viral replication, there is emerging evidence that SARS-CoV-2 co-opts and recruits additional host proteases for transmissibility.^31,32^ It is however unknown whether there is placental expression of host proteases (such as TMPRSS2 and others) necessary for cleavage of the S protein and receptor priming. On the other hand, placental barrier damage caused by severe maternal hypoxemia in women with COVID-19 infection could also be one potential way through which SARS-CoV-2 can cause intrauterine infection. There is an urgent need to further investigate the possibility of vertical transmission of SARS-CoV-2 [R,R].

The second case of maternal death that was reported by Karami et al underwent autopsy and histopathologic evaluation of paraffin embedded lung tissue that showed alveolar spaces with focal hyaline membrane, pneumocyte proliferation, and metaplastic changes.^21^ There was evidence of viral pneumonia (viral cytopathic effect including multinucleation and nuclear atypia and a mild increase in alveolar wall thickness). These findings are comparable to what have been observed in nonpregnant cases.^33^ A recent case-series reported on maternal death of seven pregnant women presenting with severe COVID-19 during the latter second and third trimester over a 30-day interval in Iran.^34^ All cases received a three-drug regimen, which included oseltamivir 75 mg PO every 12 hours for 5 days, hydroxychloroquine sulfate 400 mg PO daily or chloroquine sulfate 1000 mg tablet PO as a single dose and lopinavir/ritonavir 400/100 mg PO every 12 hours for 5 days. There were two key distinguishing features from these cases. First, the average maternal age of this series (36.7 ± 7.3 years versus 30.3 ± 3.6 years) was higher than others. Second, none of the pregnant patients had pre-existing co-morbidities, such as hypertension, cardiovascular disease and asthma.^34^ The authors believed that the COVID-19-infected pregnant patients did not receive suboptimal care and raised concern regarding the potential for maternal death among pregnant women diagnosed with COVID-19 in the latter trimester. Notably, the authors did not provide details of cause(s) of death.^34^

This systematic review had several strengths. We undertook a meticulous process in identifying duplicate reporting: (i) case reports from China were excluded, (ii) when a hospital had published their cases more than once, only the paper with the biggest data or published on the later date was included, (iii) we were able to identify the duplicates from our own publication,^11^ which is the biggest cohort series with available pregnancy outcome data from China and (iv) when necessary we contacted the corresponding authors using their native language, i.e. Chinese and Spanish, thus increasing the chance of receiving a response from them. We did not apply any language restrictions in order to capture a global picture of the impact of COVID-19 on pregnancy and perinatal outcomes and most importantly we had the capability to ensure that data from China could be accurately interpreted and recorded. There were several major limitations. We were unable to include the second biggest cohort series with available pregnancy outcome data from China because the data were pooled from a national registry.^35^ Despite contacting the corresponding author directly, it was not possible to identify the actual sources of cases. All 68 pregnant patients with pregnancy outcome data were cases from Wuhan, and the authors did not make reference to previously published cases. In our publication we included 77 cases from Wuhan and we had strong reasons to believe that there was significant overlap between the two series, and therefore we regrettably had to exclude the data from Chen et al in order to avoid inflating the number of COVID-19-infected pregnant cases in China. We also learned that COVID-19-infected pregnant cases could have been transferred to other hospitals, which made it difficult to determine duplicate reporting as cases could have been reported by both the admitting and receiving hospitals. We decided to combine laboratory-confirmed and clinically diagnosed COVID-19 cases in our analysis because both groups had similar outcomes.^11^ We believed it was important to include the clinically diagnosed cases in this review as clinically diagnosed cases had typical COVID-19 chest CT findings and significant epidemiological exposure. Only two cases of maternal death had been reported, as of April 23, 2020, in the literature. Traditionally any maternal death requires an extensive process of investigation and therefore it is not immediately reported and made public. It is highly probable that COVID-19-related maternal case fatality rate is higher but such data will only come to light at the end of the pandemic.

In conclusion, the clinical characteristics of pregnant women with COVID-19 pneumonia are similar to those of nonpregnant adults with COVID-19 pneumonia. Currently, there is no evidence that pregnant women with COVID-19 are more prone to develop severe pneumonia, in comparison to nonpregnant patients. The subject of vertical transmission of SARS-CoV-2 remains controversial and more data is needed to investigate this possibility. Lastly, in order to collect meaningful pregnancy and perinatal outcome data, we urge researchers and investigators to reference previously published cases in their publications and to record such reporting when the data of a case is being entered into a registry or several registries.

## Data Availability

This was a systematic review of published literature.

## Acknowledgments

We wish to thank Marisa Maquedano from iMaterna Foundation for assisting with the literature search.

## Appendix 1. Systematic search

(COVID-19[tw] OR COVID19[tw] OR 2019-nCoV[tw] OR nCoV-2019[tw] OR coronavirus 2019[tw] OR 2019 coronavirus[tw] OR corona virus* 2019[tw] OR 2019 corona virus*[tw] OR novel coronavirus[tw] OR novel corona virus*[tw] OR “severe acute respiratory syndrome coronavirus 2”[tw] OR coronavirus disease-19[tw] OR coronavirus disease-2019[tw] OR coronavirus disease 2019[tw] OR corona virus disease 2019[tw] OR corona virus disease-2019[tw] OR corona virus disease-19[tw] OR new coronavirus*[tw] OR new corona virus*[tw] OR SARS-CoV-2[tw] OR SARS-coronavirus 2[tw] OR “coronavirus 2”[tw] OR “spike glycoprotein, COVID-19 virus” [Supplementary Concept] OR “LAMP assay” [Supplementary Concept] OR “COVID-19 serotherapy” [Supplementary Concept] OR “COVID-19 drug treatment” [Supplementary Concept] OR “COVID-19 diagnostic testing” [Supplementary Concept] OR “COVID-19 vaccine” [Supplementary Concept] OR “severe acute respiratory syndrome coronavirus 2” [Supplementary Concept] OR (Wuhan[tiab] AND (coronavirus*[tiab] OR corona virus*[tiab])) AND (2019:2020[pdat])) AND (((((“Infant”[Mesh] OR infan*[tw] OR “Infant, Newborn”[Mesh] OR newborn*[tw] OR “Premature Birth”[tw] OR premature births[tw] OR “Infant, Premature”[Mesh] OR premature infant*[tw] OR premature bab*[tw] OR premature child*[tw] OR neonatal prematurity[tw] OR preterm birth*[tw] OR preterm infant*[tw] OR preterm bab*[tw] OR preterm child*[tw] OR pre-term birth*[tw] OR pre-term infant*[tw] OR pre-term bab*[tw] OR pre-term child*[tw] OR “Infant, Extremely Premature”[Mesh] OR extremely premature[tw] OR extremely preterm[tw] OR extremely pre-term[tw] OR “Infant, Low Birth Weight”[Mesh] OR low birth weight*[tw] OR “Infant, Very Low Birth Weight”[Mesh] OR very low birth weight*[tw] OR “Infant, Small for Gestational Age”[Mesh] OR “Infant, Postmature”[Mesh] OR postmature bab*[tw] OR postmature infant*[tw] OR postmature child*[tw] OR Perinatology[tw] OR perinatal*[tw] OR antepartum[tw] OR ante-partum[tw] OR intra-partum[tw] OR intrapartum[tw] OR Neonatology[tw] OR neonat*[tw] OR neo-nat*[tw] OR postnatal*[tw] OR post-natal*[tw] OR fetus*[tw] OR foetus*[tw] OR fetal*[tw] OR foetal*[tw] OR baby*[tw] OR babies[tw])) OR (maternal and perinatal outcomes with COVID-19 a systematic review)) OR (Pregnancy[tw] OR pregnancies[tw] OR pregnan*[tw] OR gestation*[tw] OR delivery[tw] OR deliveries[tw])))

This search is continuously updated with new terms and conditions and freely available at: https://bvcsmaquedano.wordpress.com/2020/04/17/covid-19-pregnancy/

